# Interaction-based metabolomics identifies serum modifiers of the clinical expression of Alzheimer’s disease pathology

**DOI:** 10.64898/2026.06.30.26356962

**Authors:** Pyry Helkkula, Miriam Rabl, Christopher Clark, Slavisa Stojkovic, Julius Popp

**Affiliations:** Department of Adult Psychiatry and Psychotherapy, University Hospital of Psychiatry Zurich and University of Zurich, Switzerland; European Bioinformatics Institute, European Molecular Biology Laboratory (EMBL-EBI), Cambridge, United Kingdom; Old Age Psychiatry, Department of Psychiatry, Lausanne University Hospital, Switzerland

**Keywords:** Alzheimer’s disease, Cognitive resilience, Mass spectrometry, Lipidomics

## Abstract

**Introduction:** Developing interventions for Alzheimer’s disease (AD) requires an improved understanding of mechanisms linking biological pathology to the penetrance of, and resilience to, cognitive impairment. This research identifies blood metabolites that are associated with cognitive impairment adjusting for the presence of AD pathology.

**Methods:** Mass spectrometry peaks (n=2,282) were screened using mutually conditioned models (N=154; cross-adjusting CSF-biomarker defined AD status and cognitive status). Peaks were then tested for multiplicative interactions (CSF biomarker x peak) against both endpoints using complete APOE-ε4 and albumin CSF/serum quotient data (N=140).

**Results:** The screen revealed two directional profiles: Profile I (negative with status; positive with cognition) and Profile II (positive with status; negative with cognition). Secondary interaction modeling identified four peaks with interactions significantly associated with AD, and two with cognitive impairment, including one annotated as cerebroside B.

**Discussion:** These interaction patterns indicate peripheral features acting as context-dependent modulators of AD pathology and cognitive symptom penetrance rather than global risk indicators, improving clinical stratification.

## 1 INTRODUCTION

Alzheimer’s disease (AD) is defined biologically by the accumulation of amyloid-β and pathological tau, and diagnosed via biomarkers including cerebrospinal fluid (CSF) amyloid-β42 (Aβ42) and phosphorylated tau (p-tau181) [1, 2]. Yet biological burden and functional cognitive state are only loosely coupled [2, 3, 4]. A substantial portion of older adults experience no cognitive impairment despite extensive amyloid and tau pathology, while others decline under relatively modest pathological loads [3, 4]. Understanding causes of such resilience and vulnerability to AD pathology is important for anticipating who will become symptomatic, and to developing interventions.

Systemic metabolism is one plausible source of divergence in AD symptoms. First, metabolic networks communicate with the brain across the blood–brain and blood–CSF barriers and may buffer or amplify the downstream consequences of central proteopathy. Secondly, barrier integrity itself — indexed by the CSF/serum albumin quotient (QAlb) — is altered in cognitive impairment [5, 6]. Third, circulating lipids, fatty acids and amino acids have repeatedly been associated with amyloid burden, neuropathology and cognitive trajectory, supporting the periphery as an accessible window onto these processes [7, 8].

Existing biomarkers, however, are designed to detect pathology, not to explain its variable clinical penetrance. Established AD fluid and imaging biomarkers, as well as blood-based assays such as plasma p-tau217 and the amyloid-beta (A*`β`*) 42/40 ratio, track the presence of AD pathology [9, 10]. However, they are not designed to capture the conditional modifiers that influence when and how that pathology is expressed clinically. Meanwhile, prior metabolomic studies have largely tested associations with biochemical pathology and with cognitive symptoms separately. Pathology and cognition are strongly correlated; therefore, examining associations in isolation does not allow distinguishing metabolic variance attributable to biochemical disease staging vs. overt clinical expression [7, 8].

This study therefore aims to identify blood-based metabolites that are associated with cognitive impairment adjusting for the presence of AD pathology. Using an untargeted serum mass-spectrometry pipeline (2,282 peaks) in a deeply phenotyped, CSF-characterised cohort, we applied a mutual-conditioning strategy that cross-adjusts biochemical AD status and cognitive status to isolate these two sources of variance, then tested prioritised features for multiplicative interactions with continuous CSF Aβ42 and p-tau181. The goal was to identify peripheral features that conditionally modify the clinical expression of established central pathology, and thereby improve biological and clinical stratification, rather than features that merely mirror pathology itself.

## 2 METHODS

### 2.1 Study cohort

Participants aged 49 to 88 years were recruited for an Alzheimer’s disease (AD) biomarker discovery study at the University Hospital of Lausanne, Switzerland, between 2014 and 2018. Patients were enrolled through the memory clinics of the Department of Psychiatry and the Department of Clinical Neurosciences; cognitively unimpaired control participants were recruited via targeted advertisements and word-of-mouth.

All participants underwent comprehensive clinical, neuroimaging, and neuropsychological assessments at baseline and follow-up, as previously described [11, 12]. The neuropsychological battery evaluated performance across multiple cognitive domains, including memory, language, attention, executive function, and visuospatial abilities. Global cognition was assessed using the Mini-Mental State Examination (MMSE) [13], and functional independence was evaluated using the Instrumental Activities of Daily Living (IADL) scale [14]. Collectively, the clinical examinations and neuropsychological performance indices were synthesized to determine global Clinical Dementia Rating (CDR) scores [15, 16].

Exclusion criteria consisted of a history of major psychiatric or neurological disorders (other than AD), active substance abuse, or severe, unstable systemic physical illnesses that could independently alter cognitive function or peripheral metabolic pathways. Structural neuroimaging (magnetic resonance imaging [MRI] or computed tomography [CT]) was performed on all participants at baseline to exclude individuals with confounding macrostructural cerebral pathologies (e.g., massive cortical infarcts, severe normal-pressure hydrocephalus, or intracranial masses) that could interfere with cognitive performance evaluation. A total of 154 individuals with complete baseline covariate datasets were included in the primary discovery analyses.

### 2.2 Blood and CSF collection and targeted biomarker assays

Paired venous and lumbar punctures were performed between 8:30 AM and 11:00 AM following an overnight fast. Ten to twelve milliliters of cerebrospinal fluid (CSF) were collected for analysis; initial routine protein quantification tests and cell counting were conducted immediately. The remaining CSF was immediately aliquoted (500 *`µ`*L) and frozen at −80 °C within 1 hour of collection. Serum samples were centrifuged at 4 °C, immediately aliquoted, and frozen at −80 °C no later than 1 hour post-collection until assayed, as previously described [12]. All samples were kept cooled during all preanalytical steps from the moment of collection until they were permanently frozen. Following sample preparation, CSF amyloid-beta 1-42 (*A`β`*_42_) and tau phosphorylated at threonine 181 (*p*-tau_181_) concentrations were measured using commercially available ELISA kits (Innotest®, Fujirebio Europe, Gent, Belgium). The albumin CSF/serum quotient (*Q*_alb_), used as a marker of blood-CSF barrier integrity, and the APOE genotype were determined as previously described [12].

### 2.3 Mass-spectrometric measurements

Mass-spectrometric (MS) measurements were conducted at the Metabolomics Unit of the Functional Genomics Center Zurich. Serum samples were thawed on ice, and a 50 *`µ`*L aliquot of each sample was mixed with 200 *`µ`*L of HPLC-grade methanol in a 1.5 mL microcentrifuge tube. The mixture was vortexed for 10 s and incubated for 20 min on ice. Following centrifugation (16,000 *× g*, 4 °C, 15 min), 200 *`µ`*L of the metabolite-containing supernatant was transferred to a 1 mL glass vial and stored at −20 °C until analysis. The remaining pellet was reserved for protein quantification to normalize metabolite abundances to total serum protein content.

Methanol extracts (50 *`µ`*L) were dried under a nitrogen stream, reconstituted in 20 *`µ`*L of MS-grade water, and diluted with 80 *`µ`*L of injection buffer. The mixture was vortexed and centrifuged (16,000 *× g*, 4 °C, 15 min), and 50 *`µ`*L of the supernatant was transferred to a total recovery glass vial (Waters, Milford, MA USA) for LC-MS injection. Method blanks, quality control (QC) standards, and pooled matrix samples were prepared identically to monitor instrument stability. The injection buffer comprised 90 parts acetonitrile, 9 parts methanol, and 1 part 5 M ammonium acetate.

Metabolites were separated using a nanoAcquity UPLC system equipped with a BEH Amide capillary column (150 mm *×* 50 *`µ`*m, 1.7 *`µ`*m particle size; Waters). The mobile phase gradient consisted of 5 mM ammonium acetate in water (A) and 5 mM ammonium acetate in acetonitrile (B), ramping from 5% A to 50% A over 12 min. The injection volume was 1 *`µ`*L, and the flow rate was dynamically adjusted across the gradient from 3 to 2 *`µ`*L/min. The UPLC system was coupled online to a Synapt G2Si mass spectrometer (Waters) via a nanoESI source. Data were acquired in MS^E^ mode using negative electrospray ionization across a mass range of 50 to 1200 *m*/*z* at a resolution > 20, 000 for both MS1 (molecular ion) and MS2 (fragment) scans.

Negative electrospray ionization was selected to enhance detection of deprotonated/anionic metabolites, including organic acids and acidic/polar lipid species [17, 18, 19, 20], and to improve coverage of lipid classes repeatedly implicated in Alzheimer’s disease pathophysiology [21, 22]. Untargeted processing of the raw datasets was performed using Progenesis QI software [23]. Chromatographic retention time alignment was executed against a reference map generated from the pooled QC sample, followed by automated peak picking on an aggregate ion intensity map constructed across all injections. In total, 3,760 distinct features were quantified per sample, from which 2,282 high-quality features passing standard data-completeness and quality control filtering were retained for downstream association analyses.

### 2.4 Data preprocessing and phenotypic classification

#### 2.4.1 Biochemical staging

Participants were classified according to the presence or absence of central AD pathology based on their cerebrospinal fluid (CSF) biomarker profiles. Individuals exhibiting a CSF *p*-tau_181_/*A`β`*_42_ ratio > 0.0779 were categorized as CSF positive (CSF AD+), whereas those below this threshold were categorized as CSF negative (CSF AD−). This cut-off threshold was internally established using institutional data from 120 participants, representing the fitted value that optimized group separation and maximized the Youden index in a receiver operating characteristic (ROC) analysis predicting CDR categories [12], in alignment with validation literature [11].

### 2.4.1 Clinical cognitive categorization

Concurrently, participants were functionally stratified into operational clinical phenotypes based on their baseline global CDR scores. Individuals presenting with a CDR score of 0 were classified as cognitively normal (CN) unimpaired. Individuals presenting with a CDR score > 0 were classified as cognitively impaired, a designation spanning subjects meeting criteria for either mild cognitive impairment (MCI; CDR = 0.5) or mild dementia (CDR = 1.0).

### 2.4.2 Variable preprocessing

For the primary downstream association models in the metabolite screening workflow (see Figure 1), all continuous variables, including mass spectrometry (MS) peak intensities, underwent inverse-rank normalization and subsequent standardization (*z*-scoring via mean-centering and scaling to unit variance). Raw, untransformed continuous data were retained exclusively for the baseline cohort descriptive statistics presented in Table 1.

**FIGURE 1.**
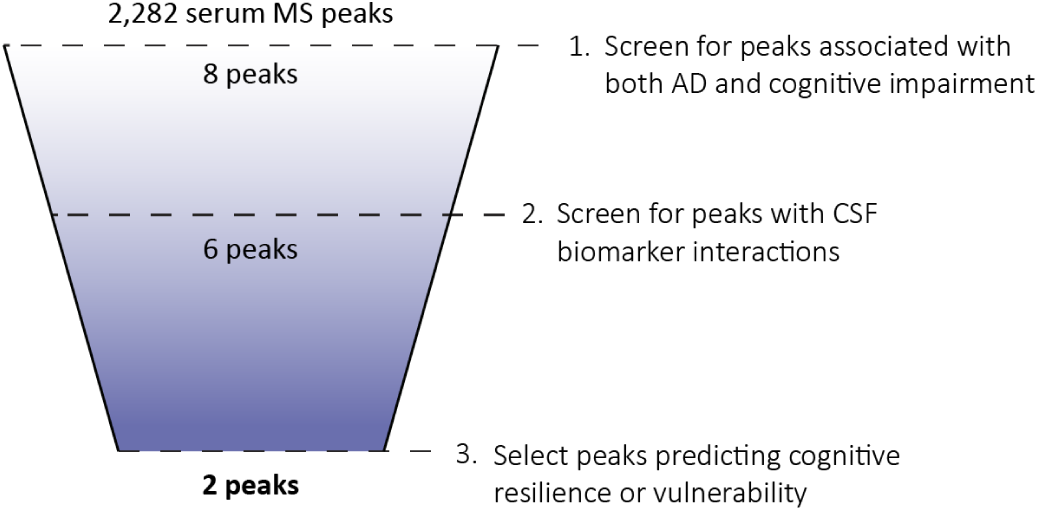
Schematic flowchart of the peripheral metabolite screening and clinical stratification workflow. An initial set of 2,282 untargeted serum mass spectrometry (MS) peaks (*N* = 154) was filtered using mutually conditioned models cross-adjusting for AD status, cognitive status, age, sex, and BMI, yielding eight peaks with a non-null posterior probability > 0.80 (local FDR < 0.20). These features were evaluated for multiplicative interactions (CSF biomarker *×* MS peak) against both endpoints in a covariate-complete subcohort (*N* = 140) adjusting for APOE-*`ɛ`*4 allele counts and blood-brain barrier integrity (*Q*_alb_). Six peaks displayed significant two-tailed interaction effects (*P* < 0.05) involving either CSF biomarker and either endpoint. Integrating structural annotation via the Human Metabolome Database (HMDB) and our established validation protocol (see Supplementary Methods) with an analysis of the interaction term signs resolved two putative metabolic markers of cognitive resilience or vulnerability.

**TABLE 1.** Baseline demographic, clinical, and fluid biomarker profiles of the study cohort. Data are stratified by biochemical AD status (AD) and clinical cognitive impairment (CI). *APOE and QAlb statistics are based on 140 samples instead of 154.

| Demographic / clinical Profile | Overall | Biochemical AD status |  | Clinical cognitive status |  | P-values |
| --- | --- | --- | --- | --- | --- | --- |
|  |  | AD– | AD+ | Normal | Impaired |  |
| Sex, female (%) | 95 (61.7%) | 62 (64.6%) | 33 (56.9%) | 46 (65.7%) | 49 (58.3%) | AD: 0.44<br>CI: 0.44 |
| Cognitive impairment (%) | 84 (54.5%) | 39 (40.6%) | 45 (77.6%) | 0 (0.0%) | 84 (100.0%) | AD: 1.74E-5<br>CI: N/A |
| Age, years ($\mu \pm$ SD) | 71.5 ± 7.59 | 69.6 ± 7.85 | 74.8 ± 5.90 | 68.4 ± 7.85 | 74.1 ± 6.32 | AD: 2.68E-5<br>CI: 1.52E-6 |
| Education, years ($\mu \pm$ SD) | 12.4 ± 2.85 | 12.4 ± 2.96 | 12.5 ± 2.69 | 12.5 ± 2.67 | 12.3 ± 3.02 | AD: 0.71<br>CI: 0.67 |
| BMI ($\mu \pm$ SD) | 25.0 ± 3.79 | 25.6 ± 4.09 | 23.9 ± 2.99 | 25.2 ± 4.06 | 24.8 ± 3.57 | AD: 7.00E-3<br>CI: 0.52 |
| QAlb ($\mu \pm$ SD)* | 6.31 ± 2.69 | 6.25 ± 2.48 | 6.41 ± 3.05 | 5.56 ± 1.88 | 6.98 ± 3.11 | AD: 0.73<br>CI: 1.60E-3 |
| APOE ε4 allele count (%)* | 52 (18.6%) | 16 (9.1%) | 36 (34.6%) | 17 (12.9%) | 35 (23.6%) | AD: 2.63E-7<br>CI: 0.03 |
| CSF Aβ42, pg/mL ($\mu \pm$ SD) | 889.4 ± 306.77 | 1048.9 ± 254.67 | 619.5 ± 166.48 | 1013.6 ± 283.63 | 778.6 ± 284.89 | AD: 3.16E-20<br>CI: 2.85E-6 |
| CSF p-tau181, pg/mL ($\mu \pm$ SD) | 65.3 ± 34.34 | 49.3 ± 15.30 | 92.4 ± 40.31 | 56.4 ± 22.84 | 73.3 ± 40.58 | AD: 1.53E-15<br>CI: 3.48E-3 |

### 2.5 Statistical analysis

#### 2.5.1 Cohort descriptive statistics

Demographic and clinical characteristics were compared across clinical groups (CSF AD– vs. CSF AD+; cognitively normal vs. cognitively impaired). Continuous variables were analyzed using Welch’s *t*-tests, and categorical variables were evaluated via chi-squared tests. Baseline data are presented as mean *±* standard deviation unless otherwise specified.

### 2.5.2 Metabolite discovery screen

To identify peripheral metabolic features associated with Alzheimer’s disease (AD) pathophysiology and clinical presentation, an untargeted screen was performed across 2,282 mass spectrometry (MS) peaks in the full cohort (*N* = 154). Generalized linear models (GLMs) with a logit link function were constructed using a mutual conditioning approach designed to achieve phenotypic orthogonalization. To isolate the independent variance of each MS peak attributable to biochemical staging versus clinical symptoms, models were cross-adjusted in pairs:

1. When evaluating the association between an MS peak and biochemical AD status, clinical cognitive impairment was included as a mandatory predictor covariate.
2. Conversely, when evaluating the association between an MS peak and cognitive impairment, biochemical AD status was included as a predictor covariate.

### 2.5.3 CSF biomarker interaction screen

To determine whether prioritized peripheral metabolic networks conditionally moderate or gate central neurodegenerative pathology, multiplicative interaction testing was conducted. This secondary analysis evaluated candidate MS peaks for explicit cross-talk with central cerebrospinal fluid (CSF) biomarkers (CSF *A`β`*_42_ or *p*-tau_181_).

Modeling was restricted to participants with complete covariate records (*N* = 140). Fully adjusted GLMs were deployed to test the significance of the multiplicative interaction term (CSF biomarker *×* MS peak) against both primary dependent variables: biochemical AD status and clinical cognitive impairment. Models included standard baseline covariates (either CSF-biomarker defined AD or cognitive status, age, sex, BMI) alongside explicit adjustments for APOE-*`ɛ`*4 allele counts and the cerebrospinal fluid-to-serum albumin quotient (*Q*_alb_) to control for blood-brain barrier integrity. Interaction effects were considered statistically significant at a two-tailed *P* < 0.05.

For peaks yielding significant interactions, average marginal effects (AMEs) were calculated and plotted across the continuous ranges of the corresponding moderators to resolve the directional nature of the conditional cross-talk. These post-estimation analyses quantified: (1) the marginal effect of serum MS peak intensities on the phenotype probability (*`∂`P*/*`∂`*MS peak) across the continuous CSF biomarker spectra, and (2) the marginal effect of central CSF biomarkers (*`∂`P*/*`∂`*CSF biomarker) across the continuous MS peak intensity spectrum.

### 2.5.4 Software and packages

All statistical analyses were executed using Python (version 3.12.2). Modeling and data configurations were implemented leveraging the following packages: statsmodels [24], scipy [25], and patsy [26]. Detected ions were identified based on accurate mass, detected adduct patterns and isotope patterns by comparing with entries in the Human Metabolome Data Base (HMDB). A mass accuracy tolerance of 10 ppm was set for the searches. Fragmentation patterns were considered for the identifications of metabolites. For details on the protocol for annotating the MS peaks and assigning Schymanski levels, see supplementary details in Section 6.3.

All discovery models were adjusted for baseline age, sex, and body mass index (BMI). Features displaying a non-null posterior probability > 0.80 (i.e., local FDR < 0.20) across both models were prioritized for downstream analyses. This empirical Bayesian framework was selected over classical family-wise error rate (e.g., Bonferroni) or global *q*-value corrections to preserve statistical power amid the highly correlated feature architectures inherent to untargeted metabolomics. While traditional penalization methods treat features as isolated, independent hypotheses, thereby inflating Type II error rates, the local FDR approach leverages the empirical distribution of the entire dataset to calculate the post-data posterior probability of a non-null effect. Enforcing a concordant posterior threshold across both distinct models ensures strict internal consistency while retaining co-regulated biological pathways.

## 3 RESULTS

### 3.1 Study participants

Baseline demographic, clinical, and biochemical characteristics of the overall study cohort (*N* = 154), stratified by both cerebrospinal fluid (CSF) Alzheimer’s disease (AD) biomarker status and clinical cognitive status, are detailed in Table 1. When stratified by central biochemical staging, individuals in the CSF AD+ group (*n* = 58) were significantly older (74.8 *±* 5.90 vs. 69.6 *±* 7.85 years; *P* = 2.68 *×* 10^−5^) and exhibited a significantly higher prevalence of cognitive impairment (77.6% vs. 40.6%; *P* = 1.74 *×* 10^−5^) than those in the CSF AD− group (*n* = 96). The CSF AD+ group also displayed a significantly lower body mass index (BMI; 23.9 *±* 2.99 vs. 25.6 *±* 4.09; *P* = 7.00 *×* 10^−3^) and a markedly higher frequency of the APOE *`ɛ`*4 allele (34.6% vs. 9.1%; *P* = 2.63 *×* 10^−7^). As dictated by the classification criteria, the CSF AD+ cohort demonstrated heavily altered central biomarker profiles, characterized by significantly lower CSF *A`β`*_42_ levels (619.5 *±* 166.48 vs. 1048.9 *±* 254.67 pg/mL; *P* = 3.16 *×* 10^−^_20_) and significantly elevated CSF *p*-tau_181_ levels (92.4 *±* 40.31 vs. 49.3 *±* 15.30 pg/mL; *P* = 1.53 *×* 10^−15^). No significant differences between biochemical strata were observed for sex distribution (*P* = 0.44), years of education (*P* = 0.71), or the CSF/serum albumin quotient (QAlb; *P* = 0.73). Concurrently, when stratified by functional clinical phenotype, cognitively impaired individuals (*n* = 84) were significantly older than their cognitively normal peers (*n* = 70; 74.1 *±* 6.32 vs. 68.4 *±* 7.85 years; *P* = 1.52 *×* 10^−6^). Cognitively impaired participants also demonstrated a significantly higher QAlb (6.98 *±* 3.11 vs. 5.56 *±* 1.88; *P* = 1.60 *×* 10^−3^) and an enrichment of the APOE-*`ɛ`*4 allele (23.6% vs. 12.9%; *P* = 0.03). With respect to central biomarkers, the cognitively impaired group displayed significantly depressed CSF *A`β`*_42_ levels (778.6 *±* 284.89 vs. 1013.6 *±* 283.63 pg/mL; *P* = 2.85 *×* 10^−6^) alongside significantly elevated CSF *p*-tau_181_ concentrations (73.3 *±* 40.58 vs. 56.4 *±* 22.84 pg/mL; *P* = 3.48 *×* 10^−3^). No significant variations across cognitive groups were found for sex distribution (*P* = 0.44), educational attainment (*P* = 0.67), or BMI (*P* = 0.52).

### 3.2 Metabolite discovery screen and phenotypic profiles

The initial untargeted screening of 2,282 serum mass spectrometry (MS) peaks across the full cohort (*N* = 154) utilized a mutual conditioning approach to isolate feature variance related to biochemical disease staging from clinical symptoms. Applying a selection threshold requiring a non-null posterior probability (*`π`*_1_) > 0.80 (local FDR < 0.20) simultaneously across both discovery models isolated eight prioritized features. Based on the valence of their mutually conditioned regression coefficients (*`β`*), these prioritized peaks fell into two distinct, mutually exclusive directional configurations designated as Profile I (*n* = 5 peaks) and Profile II (*n* = 3 peaks). Complete association statistics for all discovery models are available in Section 6.2.

Features assigned to Profile I were characterized by a negative association with biochemical AD status (*`β`* < 0) paired with a concurrent positive association with clinical cognitive impairment (*`β`* > 0). This configuration comprised peaks m/z 115.05620, m/z 181.95135, m/z 186.06522, m/z 247.15713, and m/z 708.53613. Conversely, features assigned to Profile II demonstrated the inverse configuration, presenting a positive association with biochemical AD status (*`β`* > 0) paired with a concurrent negative association with clinical cognitive impairment (*`β`* < 0). This configuration encompassed peaks m/z 302.04441, m/z 336.01697, and m/z 451.07910.

### 3.3 CSF biomarker interaction screen

To determine whether the prioritized peripheral metabolic features conditionally modify or moderate central AD pathology, secondary multiplicative interaction screens (CSF biomarker *×* MS peak) were performed in the covariate-complete subcohort (*N* = 140). These fully adjusted interaction models incorporated baseline age, sex, and BMI alongside explicit adjustments for APOE-*`ɛ`*4 allele counts and the cerebrospinal fluid-to-serum albumin quotient (QAlb) to control for blood-brain barrier integrity. Six of the eight discovery peaks demonstrated statistically significant multiplicative interaction terms (*P* < 0.05) against either biochemical AD status or clinical cognitive impairment, as detailed in Table 2.

**TABLE 2.**
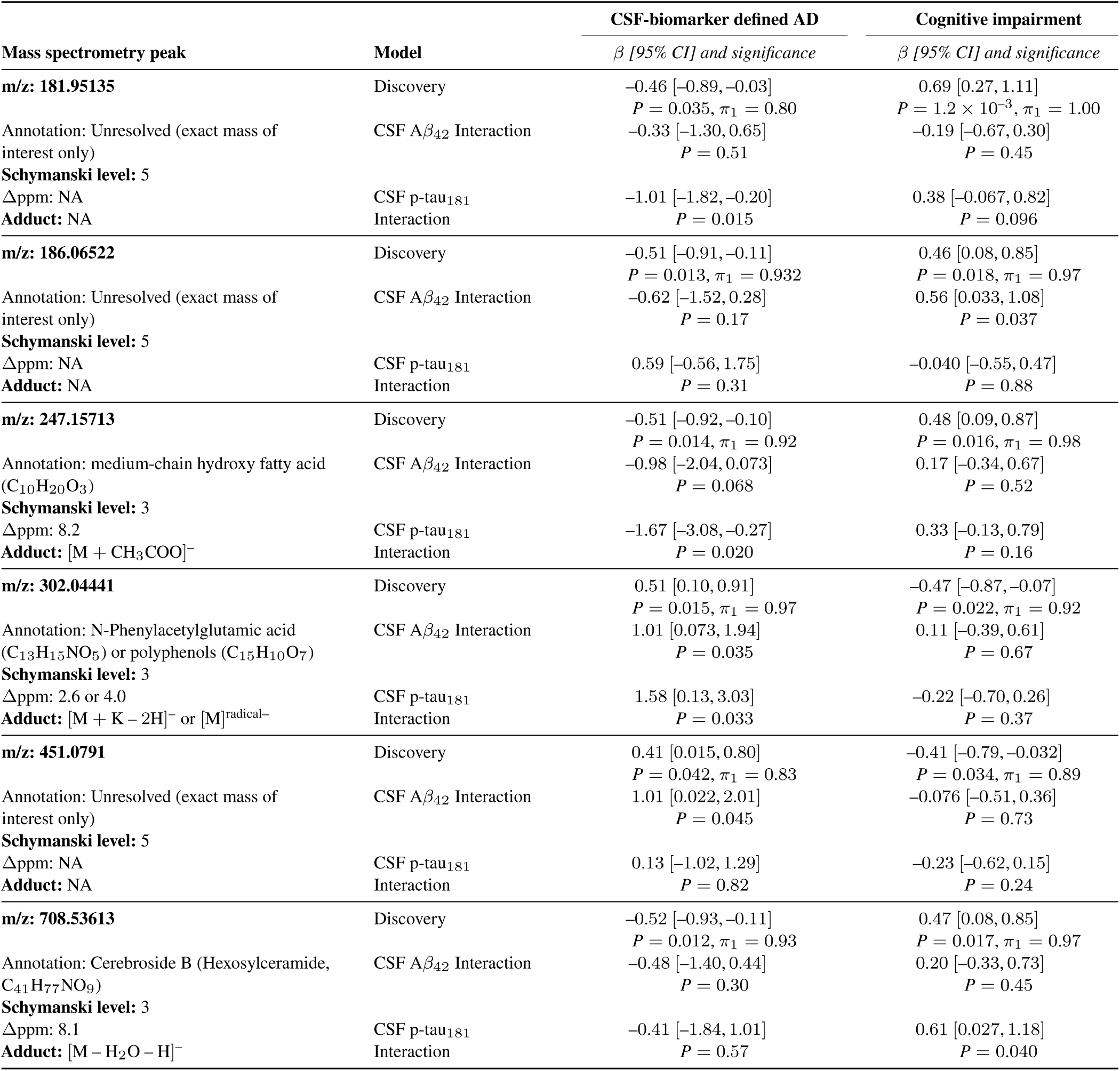
Multivariable discovery and secondary cerebrospinal fluid biomarker interaction modeling parameters for prioritized serum metabolic features. Generalized linear model parameters are presented as regression coefficients (*`β`*) with 95% CIs and significance (*P*) values. Secondary interaction models (*N* = 140) evaluate multiplicative terms (CSF biomarker*×*MS peak) adjusted for baseline demographics, APOE-*`ɛ`*4 allele counts, and blood-brain barrier integrity (*Q*_alb_). Structural chemical annotations and validation tiers are assigned based on established Schymanski levels.

Among the candidates corresponding to Profile I, two features exhibited significant interaction effects with central pathology specifically when predicting biochemical AD status. m/z 181.95135 demonstrated a significant multiplicative interaction with CSF *p*-tau_181_ for the prediction of AD status (*`β`* = –1.01, 95% CI: [–1.82, –0.20], *P* = 0.015). Similarly, m/z 247.15713 (annotated as a medium-chain hydroxy fatty acid) displayed a significant interaction term with CSF *p*-tau_181_ against the AD status endpoint (*`β`* = –1.67, 95% CI: [–3.08, –0.27], *P* = 0.020). In contrast, the remaining two interacting Profile I features demonstrated significant conditional effects when predicting clinical cognitive impairment rather than biochemical staging. m/z 186.06522 exhibited a significant multiplicative interaction with CSF *A`β`*_42_ when evaluating cognitive impairment (*`β`* = 0.56, 95% CI: [0.033, 1.08], *P* = 0.037), while m/z 708.53613 (annotated as Cerebroside B) demonstrated a significant interaction with CSF *p*-tau_181_ against the same cognitive endpoint (*`β`* = 0.61, 95% CI: [0.027, 1.18], *P* = 0.040).

For candidates categorized within Profile II, interaction modeling revealed significant cross-talk restricted exclusively to the prediction of biochemical AD status. m/z 302.04441 (annotated as N-phenylacetylglutamic acid or polyphenols) demonstrated significant multiplicative interactions across both central biomarker dimensions, showing significant cross-talk with both CSF *A`β`*_42_ (*`β`* = 1.01, 95% CI: [0.073, 1.94], *P* = 0.035) and CSF *p*-tau_181_ (*`β`* = 1.58, 95% CI: [0.13, 3.03], *P* = 0.033). Finally, m/z 451.0791 demonstrated a significant interaction effect exclusively with CSF *A`β`*_42_ when predicting biochemical AD status (*`β`* = 1.01, 95% CI: [0.022, 2.01], *P* = 0.045). None of the Profile II features displayed significant multiplicative interactions with central pathology when predicting clinical cognitive impairment.

### 3.3.1 Gating of clinical cognitive impairment risk

Post-estimation average marginal effects (AME) analysis mapped on the absolute probability scale via the multi-variable delta method resolved the conditional landscapes of the features displaying significant interaction terms against functional cognitive impairment (Figure 2). Models were explicitly adjusted for baseline CSF-biomarker defined AD status, age, sex, BMI, *Q*_alb_, and APOE-*`ɛ`*4 allele counts.

**FIGURE 2.**
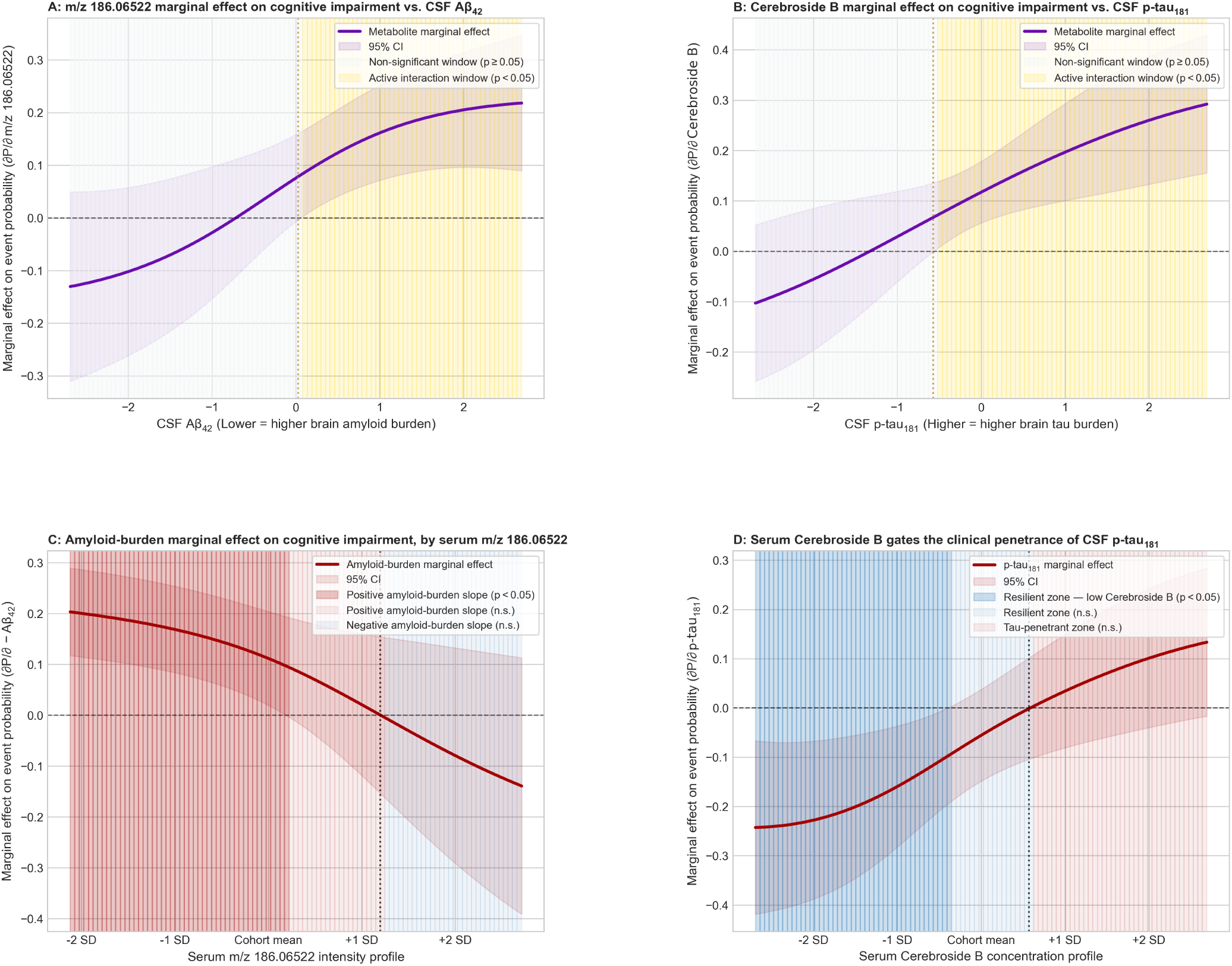
Conditional average marginal effects (AME) of serum m/z 186.06522 and Cerebroside B on cognitive-impairment probability across the CSF pathology axis. AME plots on the absolute probability scale derived via the multivariable delta method. Solid curves are point-wise marginal effects with 95% Wald confidence intervals (shaded bands). In (A) and (B), gold blocks mark active interaction windows where the confidence interval excludes zero (*P* < 0.05) and grey blocks mark non-significant windows (*P*≥0.05). In (C) and (D), shading denotes the sign of the pathology marginal effect — a positive slope (more pathology → higher impairment probability) versus a non-positive slope — solid where *P* < 0.05 and faint where *P*≥0.05; the dotted vertical line marks the sign change. All models are adjusted for baseline CSF-biomarker defined AD status, age, sex, BMI, QAlb, and APOE-ε4 allele count. **(A)** Marginal effect of serum *m*/*z* = 186.06522 (*`∂`P*/*`∂`m*/*z* 186.06522) across the CSF *A`β`*_42_ spectrum (lower values indicate higher brain amyloid burden). **(B)** Marginal effect of serum Cerebroside B (*`∂`P*/*`∂`*Cerebroside B) across the CSF *p*-tau_181_ spectrum. **(C)** Amyloid-burden marginal effect (∂P/∂−Aβ42) across the serum *m*/*z* = 186.06522 intensity range (*±*2 SD). **(D)** Tau marginal effect (*`∂`P*/*`∂`p*-tau181) across the serum Cerebroside B concentration range (*±*2 SD); the slope is negative or non-significant at low Cerebroside B (resilient profile) and significantly positive at high Cerebroside B (penetrant profile), defining a crossover interaction.

For the unresolved feature m/z 186.06522, the marginal effect on cognitive-impairment probability (*`∂`P*/*`∂`m*/*z* 186.06522) varied across the continuous CSF *A`β`*_42_ axis, with a significant active interaction window where the 95% confidence band excluded zero (Figure 2-A). The reciprocal amyloid-burden profile (∂P/∂−Aβ42) across the ±2 SD metabolite range is shown in (Figure 2-C). Because this feature remains structurally unannotated, we describe its amyloid-conditioned behaviour without assigning a directional resilience or vulnerability label.

Serum Cerebroside B (m/z 708.53613, a hexosylceramide) modified the clinical penetrance of tau pathology in a crossover fashion. The marginal effect of Cerebroside B on cognitive-impairment probability increased across the CSF p-tau181 axis, becoming a significant positive contributor under elevated tau burden (Figure 2-B). The marginal effect of CSF p-tau181 on impairment probability across the serum Cerebroside B range (±2 SD) showed that this slope changed sign with metabolite level 2-D): at low serum Cerebroside B the tau–impairment slope was negative or non-significant, whereas at high serum Cerebroside B it was significantly positive. Equivalently, among individuals with elevated tau pathology, high serum Cerebroside B accompanied a high probability of cognitive impairment (a penetrant, vulnerable profile), whereas low serum Cerebroside B accompanied preserved cognition despite comparable tau burden (a resilient profile). Cerebroside B therefore behaves as a modifier of the clinical penetrance of tau rather than as a uniform risk marker.

### 3.3.2 Modulation of CSF-biomarker defined AD risk

Four prioritized features demonstrated multiplicative interactions restricted exclusively to the prediction of biochemical AD status. For m/z 247.15713 (annotated as a medium-chain hydroxy fatty acid, C_10_H_20_O_3_), AME mapping on the real-scale probability matrix revealed a biphasic conditional profile along the tau pathology axis (Figure 3), adjusting for baseline cognitive status, age, sex, BMI, *Q*_alb_, and APOE-*`ɛ`*4 allele counts. The marginal effect of the feature on AD probability (*`∂`P*/*`∂`m*/*z* 247.15713) transitioned from a non-significant positive trend at low CSF *p*-tau_181_ to a significant negative association at high tau burden (Figure 3-A). The reciprocal profile (∂P/∂p-tau181 across the metabolite range; Figure 3-B) showed that the positive tau–AD-status slope present at low-to-average metabolite intensities was progressively attenuated as the feature accumulated, becoming non-significant above approximately +1.2 SD — i.e., higher peripheral intensities uncoupled tau burden from further change in AD-status probability.

**FIGURE 3.**
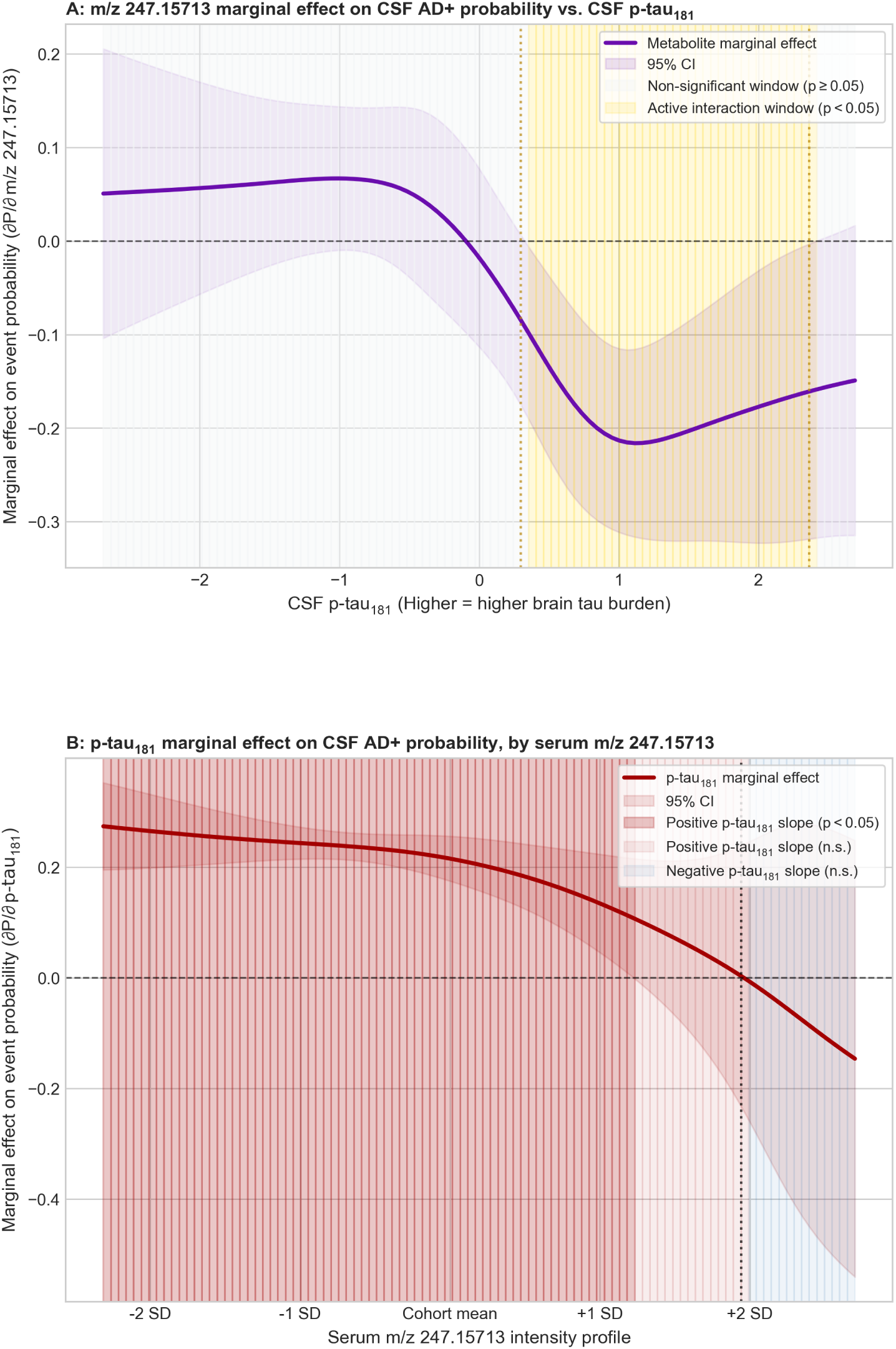
Conditional average marginal effects of serum m/z 247.15713 on CSF AD-status probability across the tau axis. AME plots on the absolute probability scale (multivariable delta method); solid curves are point-wise marginal effects with 95% Wald confidence intervals. In (A), gold blocks mark active interaction windows (CI excludes zero, p < 0.05) and grey blocks non-significant windows (*P*≥0.05). In (B), shading denotes the sign of the p-tau181 marginal effect (positive vs negative slope on AD-status probability), solid where p < 0.05 and faint where p ≥ 0.05; the dotted vertical line marks the sign change. Models are adjusted for baseline cognitive status, age, sex, BMI, QAlb, and *APOE `ε`*4 allele count. **(A)** Marginal effect of serum *m*/*z* = 247.15713 intensity on CSF-biomarker defined AD probability (*`∂`P*/*`∂`m*/*z* 247.15713) across continuous CSF p-tau181, showing a biphasic transition from a positive to a negative association. **(B)** Marginal effect of CSF *p*-tau181 (*`∂`P*/*`∂`p*-tau181) across the serum *m*/*z* = 247.15713 concentration range (*±*2 SD); the positive tau slope at lower intensities is attenuated to non-significance above +1.2 SD.

Extended post-estimation AME mapping for the remaining candidate features further resolved how peripheral metabolic variations alter absolute disease probability across continuous central pathology axes (detailed probability matrices and calculus frameworks are provided in the Supplementary Materials, see Section 6). Within the Profile I classifications, m/z 181.95135 showed significant marginal effects (*`∂`P*/*`∂`m*/*z* 181.95135) within discrete windows along the tau axis, where increasing metabolite intensity attenuated the tau-associated slope on CSF AD+ probability (Figure S1-A,C). m/z 451.0791 showed a significant marginal contribution localized around the cohort mean of amyloid burden (Figure S1-B,D). For the Profile II feature m/z 302.04441, interaction modelling indicated significant marginal contributions across broad ranges of both CSF biomarkers (Figure S2-A,B). Reciprocal profiles showed that increasing metabolite intensity attenuated the amyloid-associated slope while accentuating the tau-associated slope on CSF AD+ probability (Figure S2-C,D).

## 4 DISCUSSION

This study sought to identify blood-based metabolites that explain the residual variance in cognitive impairment once the status of central Alzheimer’s disease (AD) pathology is accounted for. An accessible, serum-based readout of penetrance or resilience would add prognostic information orthogonal to existing assays (e.g., plasma p-tau217 and the Aβ42/40 ratio) and could expose modifiable peripheral mechanisms. Using a mutual-conditioning screen to orthogonalise biochemical staging from cognitive status, followed by multiplicative interaction modelling against continuous CSF biomarkers, the analysis identified two serum features that conditionally modified cognitive impairment (Figure 2) and four that conditionally modified biochemical AD status (Figures 3, S1, S2).

The most fully resolved candidate, the hexosylceramide annotated as Cerebroside B (m/z 708.53613), showed a crossover interaction with CSF p-tau181 in predicting cognitive impairment, a pattern consistent with modification of clinical penetrance. Among individuals with elevated tau burden, high serum Cerebroside B accompanied a high probability of impairment (a penetrant, vulnerable profile), whereas low serum Cerebroside B accompanied preserved cognition despite comparable pathology (a resilient profile).

Existing sphingolipid literature links this lipid family to dementia, though the most direct predictive evidence concerns ceramides specifically. Elevated serum and plasma ceramides predict incident AD, memory decline and hippocampal atrophy [27, 28], and the plasma sphingolipidome is altered early in the disease [29]. Mechanistically, ceramide accumulation has been proposed to inhibit the insulin/PI3K–Akt cascade and thereby disinhibit glycogen synthase kinase-3β (GSK-3β), a kinase that hyperphosphorylates tau, promotes BACE1 transcription and inactivates Nrf2; through these and related effects on lipid-raft organisation and mitochondrial function, ceramide-pathway lipids could plausibly contribute to the conversion of established pathology into cognitive deficit [30]. Cell-type-resolved work further shows that neurons are comparatively sensitive to sphingolipid toxicity and that flux between ceramide and its hexosyl derivatives is tightly and differentially regulated across CNS cell types [31]. However, this mechanistic literature concerns ceramides specifically, whereas our feature is a hexosylceramide. Extending the ceramide–tau argument to it assumes that the regulated interconversion between ceramides and their glycosylated derivatives couples the two pools, which remains to be established.

The present results provide preliminary evidence of a conditional, tau-dependent hexosylceramide signal associated with cognitive impairment. A previous population study found plasma hexosylceramides — the class our feature belongs to — were not consistently altered across dementias, with reductions mainly in behavioural-variant frontotemporal dementia [32]. Notably, that null-to-negative main-effect result is for the same lipid class as our feature; our finding is a conditional, tau-dependent signal that main-effect comparisons would miss, which may explain why a hexosylceramide–AD association has not previously emerged and illustrates the value of interaction-based modelling.

The second key result of this study was an interaction between cognitive impairment and a candidate amyloid-conditioned modifier, m/z 186.06522. Its marginal association with cognitive impairment varied across the continuous CSF Aβ42 axis. Conceptually, this is the amyloid-side counterpart to the resilience–vulnerability dissociation that motivates the cognitive-reserve literature, in which amyloid burden is uncoupled from cognition in a subset of individuals [4, 3]. However, this feature remains structurally unannotated (Schymanski level 5), so it cannot yet be mapped onto a specific metabolic pathway or prior report. Thus, despite statistical evidence of amyloid-conditioned behaviour, further biological interpretation is needed.

Four features conditionally modified AD status rather than cognition. m/z 247.15713, annotated as a medium-chain hydroxy fatty acid (*C*_10_*H*_20_*O*_3_), showed a biphasic tau interaction in which higher peripheral intensities progressively uncoupled tau burden from AD-status probability. Medium-chain fatty acids are alternative cerebral energy substrates that rescue bioenergetic failure and improve cognition in MCI and AD, and mitochondrial fatty-acid β-oxidation is closely tied to AD pathophysiology [33, 34], consistent with a metabolic, energy-supportive role for this feature. m/z 302.04441 — annotated as N-phenylacetylglutamic acid (phenylacetylglutamine) or, isobarically, a polyphenol — interacted with both central biomarkers. Phenylacetylglutamine is a gut-microbial co-metabolite of dietary phenylalanine that signals through adrenergic receptors and promotes mitochondrial dysfunction and cellular senescence, and has been associated with cognitive impairment and reported as elevated in AD, situating this feature within the gut–brain axis [35, 36, 37]. The remaining two features (m/z 181.95135, m/z 451.0791) are unresolved and are best regarded as hypothesis-generating leads.

The research has several limitations. The cohort is modest (N = 154; interaction subcohort N = 140), single-centre, and not externally replicated; additionally, the design is cross-sectional, so penetrance and resilience are inferred from CSF-biomarker × metabolite interactions rather than observed from longitudinal trajectories. Annotation is a further limitation: several features are unresolved (Schymanski level 5) or ambiguously annotated (m/z 302.04441), and glucosyl- and galactosylceramide isomers cannot be distinguished by this method, so “Cerebroside B” denotes a hexosylceramide species rather than a single structure. Negative-ionisation-only LC-MS restricts metabolite coverage, the dichotomous endpoint pools MCI with mild dementia, and residual confounding (diet, medication, comorbidity, fasting) and reverse causation cannot be excluded.

Future work should pursue targeted, isomer-resolved and absolutely quantified assays of Cerebroside B and the medium-chain hydroxy fatty acid, annotation of the unresolved features, replication in independent and longitudinal cohorts to test prediction of actual decline or stability, and mechanistic studies of sphingolipid–tau and gut–microbiome pathways.

## 5 CONCLUSION

In summary, an interaction-aware, phenotypically orthogonalised analysis of the serum metabolome identifies circulating features whose associations are consistent with context-dependent modification of Alzheimer’s disease penetrance and resilience, rather than with global risk. In the clearest example, the hexosylceramide annotated as Cerebroside B showed a crossover interaction with tau pathology: higher serum levels accompanied the symptomatic, vulnerable expression of tau burden, whereas lower levels accompanied a resilient profile in which comparable pathology was uncoupled from cognitive impairment. Such peripheral markers could complement pathology-mirroring blood biomarkers and improve biological and clinical stratification in AD.

## DATA AVAILABILITY STATEMENT

In addition to the supplementary data provided, code and additional data will be made available upon reasonable request.

## ETHICS DECLARATION

The study was conducted in accordance with applicable laws and regulations, including the International Conference on Harmonization, Guideline for Good Clinical Practice and the ethical principles that have their origins in the Declaration of Helsinki [https://doi.org/10.1001/jama.2013.281053]. The local ethics committee (CER-VD, Switzerland) approved this study (No. 171/2013), and all participants or their legal representatives provided written informed consent.

## AUTHOR CONTRIBUTIONS

Pyry Helkkula - Conceptualization - lead, Writing - original manuscript, Investigation, Methodology, Visualization, Formal analysis - lead, Software, Validation, Writing - Review & Editing Miriam Rabl - Investigation, Data curation, Writing - Review & Editing Christopher Clark - Investigation, Data curation, Formal analysis - supporting, Writing - Review & Editing Slavisa Stojkovic - Writing - Review & Editing Julius Popp - Conceptualization - supporting, Supervision, Resources, Funding Acquisition, Project administration, Writing - Review & Editing

## Supporting information

MS peak annotation protocol

## ACKNOWLEDGEMENTS

We gratefully acknowledge Dr Endre Laczko (Functional Genomics Center Zurich, FGCZ) for supporting the mass-spectrometry metabolomics measurements and for valuable discussions on the interpretation of the metabolomics data.

## DISCLOSURE OF ARTIFICIAL INTELLIGENCE USE

Generative artificial intelligence was used to edit the manuscript and to assist with software development. The authors take full responsibility for the analyses and content of the manuscript.

## 6 SUPPLEMENTARY MATERIALS

### 6.1 Metabolite discovery screen association statistics

Please find the detailed data on metabolite discovery in Supplementary Tables 1-3.

### 6.2 Metabolite CSF biomarker interaction screen association statistics

Please find the detailed data on metabolite CSF biomarker interaction in Supplementary Tables 4-11.

### 6.3 MS peak annotation protocol

Please find the detailed markdown file containing the MS peak annotation protocol and the Supplementary Tables 12 and 13 with quality-controlled 2,282-feature table (per-feature metadata), and candidate annotations respectively. Supplementary Tables 14-19 contain the per-target Spearman correlation vectors for the six peaks with significant CSF biomarker interactions.

### 6.4 Supplementary figures

**FIGURE S1.**
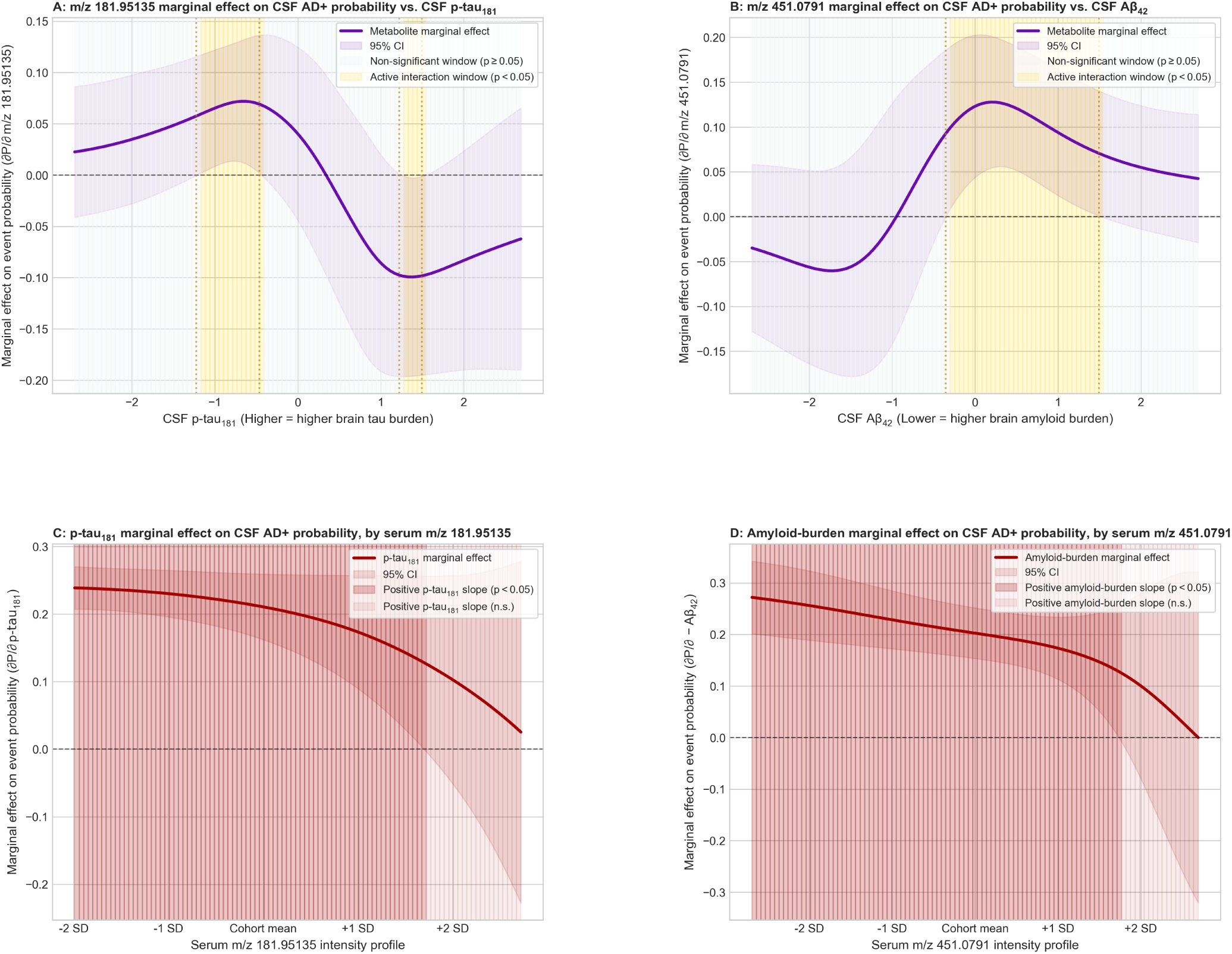
Conditional average marginal effects of serum m/z 181.95135 and m/z 451.0791 on CSF AD-status probability. AME plots on the absolute probability scale (multivariable delta method); solid curves with 95% Wald confidence intervals. In (A, B), gold/grey blocks mark active/non-significant interaction windows (p < 0.05 / p ≥ 0.05). In (C, D), shading denotes the sign of the pathology marginal effect (positive vs negative slope on AD-status probability), solid where p < 0.05 and faint where p ≥ 0.05, with a dotted line at the sign change. Both features are structurally unresolved (Schymanski level 5); their conditional effects are reported neutrally, not as resilience or vulnerability markers. Models are adjusted for baseline cognitive status, age, sex, BMI, Qalb, and APOE-ε4 allele count. (A) ∂P/∂m/z 181.95135 across CSF p-tau181. (B) ∂P/∂m/z 451.0791 across CSF Aβ42 (lower values indicate higher amyloid burden).(C) ∂P/∂p-tau181 across the serum m/z 181.95135 range (±2 SD).(D) ∂P/∂−Aβ42 (amyloid burden) across the serum m/z 451.0791 range (±2 SD).

**FIGURE S2.**
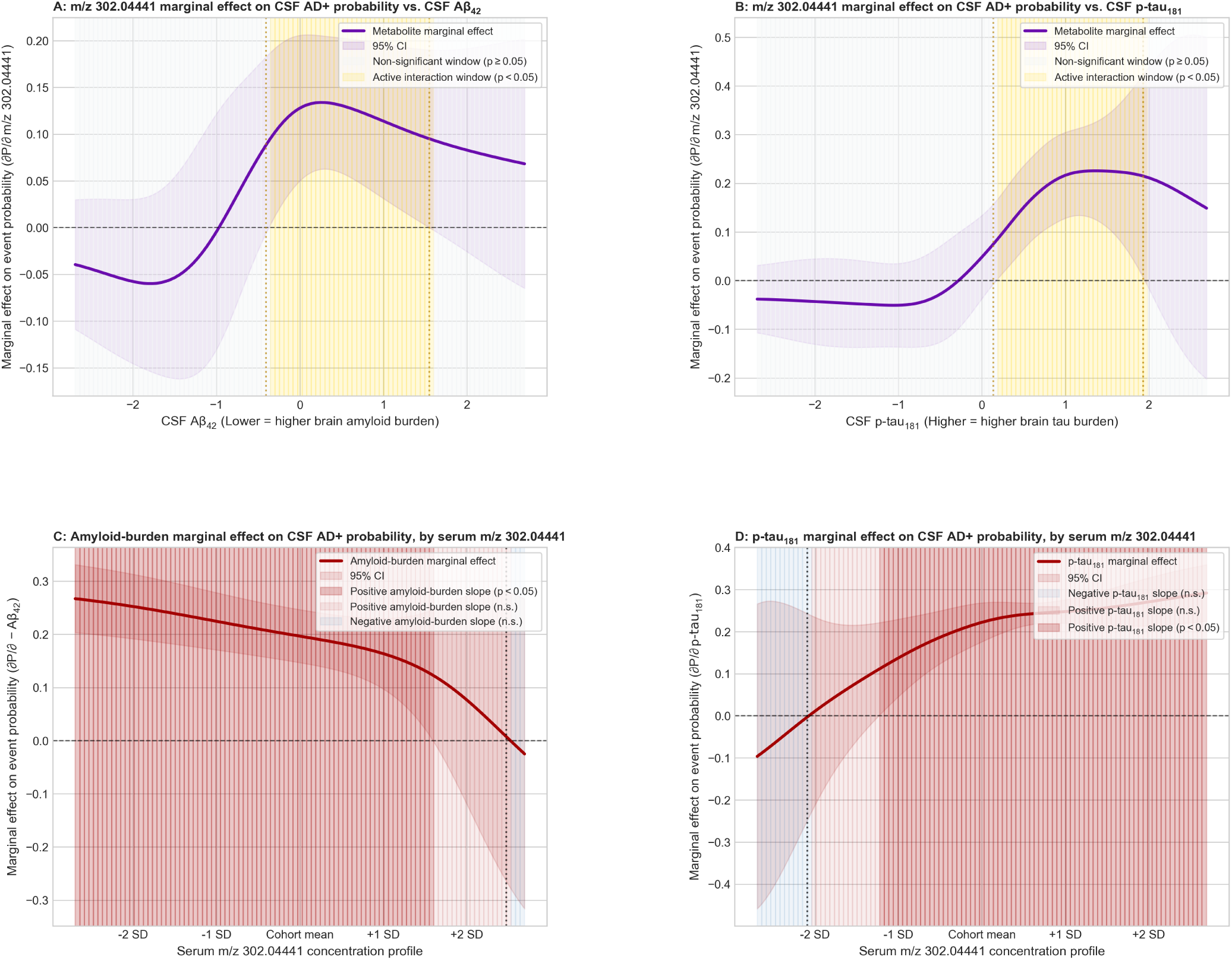
Conditional average marginal effects of serum m/z 302.04441 on CSF AD-status probability across both central biomarkers. m/z 302.04441 is annotated as N-phenylacetylglutamic acid or a polyphenol; these candidates are isobaric and were not resolved. AME plots on the absolute probability scale (multivariable delta method); solid curves with 95% Wald confidence intervals. In (A, B), gold/grey blocks mark active/non-significant interaction windows. In (C, D), shading denotes the sign of the pathology marginal effect, solid where p < 0.05 and faint where p ≥ 0.05, with a dotted line at the sign change. Conditional effects are reported neutrally, not as resilience or vulnerability markers. Models are adjusted for baseline cognitive status, age, sex, BMI, Qalb, and APOE-ε4 allele count. (A) ∂P/∂m/z 302.04441 across CSF Aβ42 (lower values indicate higher amyloid burden).(B) ∂P/∂m/z 302.04441 across CSF p-tau181. (C) ∂P/∂−Aβ42 (amyloid burden) across the serum m/z 302.04441 range (±2 SD). (D) ∂P/∂p-tau181 across the serum m/z 302.04441 range (±2 SD).

