## Supplementary material for "Interaction-based metabolomics identifies serum modifiers of the clinical expression of Alzheimer’s disease pathology": MS peak annotation protocol: MS_peak_annotation_protocol.md

# Supplementary Methods — A generalizable, adduct-agnostic protocol for annotating peaks of interest

## Scope and design principle

This protocol assigns putative identities, with an explicit confidence level, to chromatographic-mass features of interest from the negative-mode HILIC–MSᴱ serum metabolomics dataset (Progenesis QI output). It is written to generalize to any feature in the study and to accept candidate annotations carrying **arbitrary adducts**, not only the deprotonation/water-loss adducts used by the upstream software. Generalizability to arbitrary adducts is achieved by deriving every predicted quantity from two inputs only — a candidate's neutral molecular formula (which fixes its exact monoisotopic mass and theoretical isotope pattern) and an extensible adduct dictionary (each adduct defined by a charge and an additive mass transform) — and by drawing every confirmatory measurement from data that are themselves adduct-independent (observed m/z, charge, retention time, chromatographic peak width, the per-target correlation vector, and the observed isotope distribution). No step depends on a fixed, pre-enumerated adduct list.

The protocol does not require MS/MS interpretation, authentic standards, or access to raw files; with those, the achievable confidence rises (see Stage 7).

## Experimental conditions (fixed for this dataset)

These conditions are fixed inputs to the protocol; they enter only as the adduct-admissibility table (Stage 5), the retention-time-class expectations (Stage 5), and the fragmentation caveat. For another method, replace this section and the two tables it feeds; nothing else changes.

- Ionisation: electrospray, **negative** mode. This sets the sign of every adduct transform in the adduct dictionary.
- Chromatography: hydrophilic-interaction (HILIC) on a Waters BEH Amide column; gradient of 5 mM ammonium acetate in water (A) and in acetonitrile (B), 5% A to 50% A over 12 min. Retention-time expectations follow HILIC polarity ordering (nonpolar analytes elute early at high organic, polar analytes later).
- Mobile-phase additive: **5 mM ammonium acetate**, with **no formic acid/ammonium formate, no fluoride source, and no added chloride**. Admissible negative-mode adducts are therefore [M−H]⁻ and [M+CH₃COO]⁻; [M+Cl]⁻ only from trace chloride; [M−CH₃]⁻ for quaternary-amine (choline) lipids; in-source [M−H₂O−H]⁻ for labile species. Formate and fluoride adducts are inadmissible and are down-weighted if supplied as candidate inputs. Radical molecular anions [M]•⁻ (and any direct neutral-mass = observed-m/z match) are inadmissible by default and admitted only for known electron-affinic classes, per the rule in Stage 3.
- Acquisition: data-independent **MSᴱ** (no precursor isolation). Consequently a low Fragmentation Score, especially on a low-abundance feature, is uninformative and is never used to reject a candidate; MSᴱ fragments are used only when they positively show class-diagnostic ions.
- Export processing: the Progenesis QI feature table is deisotoped/deconvolved on export. Isotopologues are therefore not present as separate features, so the isotope-pattern axis (Stage 3) is taken from the per-feature Isotope Distribution column, and the co-feature test (Stage 4) relies on adduct, in-source-fragment, and charge-state siblings rather than on M+1 features.

## Co-variation engine: precomputed correlation vector (fixed for this dataset)

The Stage 4 co-variation test requires, for the target peak, its Spearman rank correlation to every other feature across the 154 study samples. To avoid handling sample-level data, this is supplied as a **precomputed per-target correlation vector**: one file per peak to be annotated, with one row per other feature giving that feature's `m/z`, the Spearman `spearman_rho` to the target, the `p_value`, and the sample count `n` (an `intensity_median` column may be included for convenience). The correlation is computed by the analyst, on their side, over the 154 study samples in their own sample identifiers; no sample-level intensities enter the workflow. Because Spearman ρ is symmetric, a target-anchored vector (target → each candidate sibling) supplies exactly what the sibling-ion test needs; this directionality should be stated when reporting. The vector is matched to the target's predicted sibling m/z within the sibling mass tolerance.

Four practical points govern its use. First, the vector's m/z field is named `m/z` and holds numeric values directly (no prefix or label formatting), matched against the predicted sibling m/z. Second, the vector should cover the full quality-controlled feature list (here 2,282 features, matching the measurements summary export). A predicted sibling whose m/z falls on a feature that was removed during quality control (the original Progenesis export was filtered from 3,760 to 2,282 features by excluding peaks that were null-heavy across samples) is therefore scored "not assessable", not "absent". Third, retention times are **not** in the correlation vector, so the co-elution filter (Stage 4) reads each candidate sibling's retention time from the Compound Measurements summary export, which therefore remains a required input alongside each vector. Fourth, the target peak itself appears in its own vector as a row with a blank/missing correlation (its self-correlation is excluded by construction); any predicted sibling m/z that maps back to the target — i.e. resolves to a row with missing ρ — is skipped.

The Compound Measurements summary export (sample-level columns removed) supplies all per-feature metadata: m/z, charge, retention time, chromatographic peak width, observed Isotope Distribution, Maximum Abundance, and Minimum CV%.

## Fixed numeric parameters and reference tables

The following defaults are used unless stated otherwise in a given run; all are reported with the results so the workflow is exactly reproducible, and any may be adjusted for another dataset.

- Candidate-search mass tolerance: **≤10 ppm** (matching how the candidate lists are generated).
- Sibling-search mass tolerance: **≤10 ppm** (kept equal to the candidate tolerance so the two are comparable).
- Co-elution retention-time window: **±0.10 min** (about one-quarter of the median chromatographic peak width of 0.41 min in this dataset); for unusually broad or narrow peaks, use ±0.5 × the query peak's own chromatographic peak width instead.
- Confirming-sibling correlation threshold: Spearman **ρ ≥ 0.70 and p < 0.05** across the 154 study samples (for reference, the genuine co-varying network observed for the worked example peaked at ρ ≈ 0.75; at n = 154, ρ = 0.70 is highly significant, so the p filter is only a guard).
- Reliability filters: flag a feature as analytically weak (carry the weakness into the confidence statement; do not exclude) when **Minimum CV% > 30%** or when **Maximum Abundance is below the dataset median (≈2,420 for this 2,282-feature set)**.
- Isotope agreement: require the observed M+1 relative intensity to be within **±30% (relative)** of the theoretical value from the candidate formula, applied **only** when Maximum Abundance is above the dataset median; below the median the isotope pattern is too noisy to weigh.
- Stage 6 ranking: use an explicit ranked ordering of evidence rather than a single weighted sum (the axes are not on commensurable scales). Order of importance: (1) number and strength of confirmed co-eluting, co-varying sibling ions; (2) adduct admissibility under the mobile phase; (3) mass error; (4) isotope-pattern agreement; (5) retention-time-class plausibility.
- Monoisotopic masses and the adduct dictionary are fixed as given in the "Reference tables fixed before analysis" section below.

## Datapoints used, and the role of each

The protocol draws on the two Progenesis QI exports and the experimental metadata. The columns and their roles are listed below; the final column marks those that make the workflow generalizable to arbitrary adducts (i.e. computable for any adduct from formula + adduct transform).

From the Compound Measurements summary export (the quality-controlled feature table; 2,282 features remaining after null-heavy peaks were removed from the original Progenesis export; sample-level columns removed, per-feature metadata retained; a single header row). Two leftover quality-control columns may be present (`_target_mz`, the feature's own m/z used as a join key, and `_mz_difference`, the self-join residual); both are ignorable and not used by the protocol.

- Observed m/z — measured ion mass; basis of the mass-accuracy test for any predicted adduct. Generalizable.
- Charge (z) — measured charge state; used to test consistency with the proposed adduct's charge and to predict multiply-charged sibling ions. Generalizable.
- Retention time (min) — used to group ion species of one neutral molecule by co-elution; adduct-independent. Generalizable.
- Chromatographic peak width (min) — defines the retention-time tolerance for the co-elution test (Stage 4); also a peak-quality metric. Generalizable.
- Observed Isotope Distribution (e.g. "100 − 24.1 …") — compared with the theoretical pattern from the candidate formula; gives an approximate carbon count (M+1 % ÷ 1.07) and a formula-consistency check; essentially adduct-independent. Generalizable.
- Precomputed per-target correlation vector (a separate input, not a column of this export) — supplies the Spearman correlation from the target peak to every other feature, computed by the analyst over the 154 study samples; this is the engine of the co-variation (deductive) test for any predicted sibling ion of any adduct. Generalizable and central.
- Maximum Abundance — reliability weight: low-abundance features have unreliable isotope patterns and weak/ambiguous MSᴱ fragmentation, and may be detectable only as a single ion; used to temper confidence.
- Minimum CV% (across QC pool injections) — analytical-reproducibility filter; features above a CV threshold are flagged as unreliable.
- Anova (p), q Value, Max Fold Change — used only to select which peaks are "of interest" and to prioritise annotation effort; not used as identity evidence. (The Highest/Lowest Mean columns of the original export are not retained in the summary file and are not required.)

From the Compound Identifications export (candidate annotations):

- Formula — the candidate neutral molecular formula; fixes the exact monoisotopic mass and theoretical isotope pattern from which all adduct m/z and all isotopologue intensities are computed. Generalizable and central.
- Compound ID (HMDB accession) and Description — database linkage and compound class, used for the retention-time-plausibility check and for reporting.
- Adducts — the adduct the software assigned; treated as one hypothesis among many, not a restriction (Stage 2 widens this).
- Mass Error (ppm) — software mass accuracy; recomputed independently for every adduct hypothesis so that arbitrary adducts are handled identically. Generalizable.
- Isotope Similarity and Theoretical Isotope Distribution — the isotope-pattern axis; adduct-independent. Generalizable.
- Score — the software's composite identity score; optional supporting weight.
- Fragmentation Score — MSᴱ fragment-match score; used cautiously and only as positive evidence (see note on MSᴱ below).
- Accepted? — analyst-validation flag, carried through to reporting.
- Charge, m/z, Neutral mass, Retention time — keys for cross-linking a candidate to its feature in the measurements table.

From the experimental metadata and earlier analysis:

- Ion polarity (negative) — sets the sign of every adduct transform. Generalizable.
- Mobile phase (5 mM ammonium acetate; no formate, no fluoride, no added chloride) — defines the adduct-admissibility table (Stage 5): abundant species are [M−H]⁻ and [M+CH₃COO]⁻; [M+Cl]⁻ only from trace chloride; [M−CH₃]⁻ for quaternary-amine (choline) lipids; in-source [M−H₂O−H]⁻ for labile species; formate and fluoride adducts inadmissible. Adducts outside this set are not forbidden as inputs but are down-weighted.
- Column chemistry (BEH Amide HILIC) — sets retention-time expectations by analyte polarity (Stage 5).
- Acquisition (MSᴱ, data-independent) — fragmentation caveat: because precursors are not isolated, a low Fragmentation Score on a low-abundance feature is uninformative (it reflects co-elution interference and weak signal), so MSᴱ fragmentation is used only when it positively shows diagnostic ions, never to reject a candidate.
- Deisotoping/deconvolution of the exported table — because isotopologues were collapsed during export, the co-elution/co-variation test (Stage 4) is run on adduct, in-source-fragment, and charge-state siblings (which remain as separate features), while the isotope-pattern axis is taken from the per-feature Isotope Distribution column rather than by searching for a separate M+1 feature.
- Monoisotopic element masses and the adduct dictionary — the fixed reference tables that make all predictions deterministic and reproducible.

## Reference tables fixed before analysis

(1) Monoisotopic masses (Da): electron 0.000549; H 1.007825; proton (H⁺) 1.007276; ¹³C−¹²C 1.003355; C 12.000000; N 14.003074; O 15.994915; P 30.973762; S 31.972071; F 18.998403; ³⁵Cl 34.968853; Na 22.989770; K 38.963707.

(2) Adduct dictionary — the extensibility point. Each entry is a tuple (label, charge z, multiplicity n, mass transform Δ), and the predicted ion m/z for a neutral monoisotopic mass M is m/z = (n·M + Δ + z·mₑ) / |z| for negative ions, where Δ is the net added/removed atomic mass excluding the charging electrons. Initial negative-mode entries: [M−H]⁻ (z1, n1, Δ = −1.007825); [M+CH₃COO]⁻ (z1, n1, Δ = +59.013304); [M+Cl]⁻ (z1, n1, Δ = +34.968853); [M−CH₃]⁻ (z1, n1, Δ = −15.022928, choline lipids); [M−H₂O−H]⁻ (z1, n1, Δ = −19.018390, in-source); [M−2H]²⁻ (z2, n1, Δ = −2.015650); [2M−H]⁻ (z1, n2, Δ = −1.007825); [M+HCOO]⁻ and [M+F]⁻ included for completeness but flagged inadmissible for this mobile phase. **Any further adduct is supported by adding one row to this dictionary; nothing else in the protocol changes.** A candidate supplied with an arbitrary adduct is parsed through the same dictionary (or the user supplies its z, n, and Δ), so candidate inputs are not restricted to the upstream software's adduct set.

(3) Adduct-admissibility weights for this mobile phase (Stage 5), and (4) compound-class retention-time windows for BEH Amide HILIC (Stage 5), both editable per method.

## Inputs required per run

To annotate a given peak, the following are needed; the first three are shared across all peaks, the fourth is peak-specific, and the last two are supplied in the request.

1. Compound Measurements summary export (quality-controlled 2,282-feature table; per-feature metadata: m/z, charge, retention time, chromatographic peak width, observed Isotope Distribution, Maximum Abundance, Minimum CV%).
2. Compound Identifications export (candidate formulas, adducts, Isotope Similarity, Theoretical Isotope Distribution, Mass Error, Score, Fragmentation Score, Accepted?).
3. A precomputed per-target correlation vector for the peak being annotated (one row per feature: a numeric `m/z` column, plus `spearman_rho`, `p_value`, `n`; computed by the analyst over the 154 study samples) — the co-variation engine for Stage 4. Covering the full quality-controlled 2,282-feature list (the target itself appears as a blank-correlation row); predicted siblings mapping to QC-removed features are scored "not assessable".
4. A per-peak candidate list with arbitrary adducts (formula, neutral mass, adduct, adduct m/z, ppm) from a broad accurate-mass database search at ≤10 ppm.
5. The target peak's observed **m/z and retention time** (retention time is required because distinct features can share a near-identical mass; e.g. 708.5361 at 4.56 min versus 708.5302 at 5.09 min).
6. Confirmation that the Experimental conditions, Sample selection, and Fixed numeric parameters sections above apply unchanged, or the specific overrides for this run.

No other information or files are required to reproduce the annotation at the confidence level the data support; the raw MSᴱ files, authentic standards, and spectral libraries are needed only to reach Schymanski Levels 1–2.

## Stepwise protocol

**Stage 1 — Select peaks of interest and apply reliability filters.** Define the peaks to annotate from the study's statistical results (e.g. features associated with CSF-biomarker–defined AD status, with cognitive impairment, or with a significant interaction term), using the Anova (p)/q Value/Max Fold Change columns or the external association models. For each selected feature, record from the measurements table its observed m/z, charge, retention time, chromatographic peak width, Maximum Abundance, observed Isotope Distribution, and Minimum CV%. Flag features with Minimum CV% above the study's QC threshold (e.g. >30%) or with very low Maximum Abundance as analytically weak; these may still be annotated but the weakness is carried into the confidence statement.

**Stage 2 — Assemble the candidate set with arbitrary adducts.** For each peak, collect candidate (neutral formula, adduct) hypotheses from all available sources: the Progenesis Identifications rows for that feature, and any external accurate-mass searches against compound databases (e.g. HMDB, LIPID MAPS, KEGG; search more than one, as a single database's coverage gaps cause false "no candidate" outcomes) performed with a broad adduct list and a defined mass tolerance (e.g. ≤10 ppm), exactly as in the supplied candidates file. The database search must convert the observed m/z to a neutral mass *via each adduct* (for example, neutral = m/z + 1.007276 for [M−H]⁻) before matching, rather than matching database neutral masses directly to the observed m/z; a negative-mode peak whose only hit is a compound whose *neutral* mass equals the observed m/z is the signature of the latter error and of a spurious radical/neutral match. Adducts from any source are admitted, including radical and uncommon species; each is parsed through the adduct dictionary. Collapse database entries sharing one (formula, adduct) into a single hypothesis (isomers are resolved later or reported jointly). Each hypothesis carries a neutral monoisotopic mass M computed from its formula. If no database candidate survives Stage 3/5 under an admissible adduct, run a fallback molecular-formula generator at the admissible-adduct neutral mass (CHNOPS, optionally Na/K/Cl; element ranges and a ring-plus-double-bond (RDBE) constraint of 0–≈16 declared in advance; mass tolerance as for the candidate search), so the peak can still reach Level 4 (an unequivocal molecular formula) when one and only one admissible formula exists, instead of defaulting to Level 5 merely because no database entry was found.

**Stage 3 — Per-candidate consistency checks (mass, charge, isotope).** For each hypothesis, using the proposed adduct's dictionary entry: (i) compute the predicted ion m/z and the mass error in ppm against the feature's observed m/z; retain only hypotheses within the mass tolerance. (ii) Require the adduct's charge z to equal the feature's measured Charge; reject on mismatch. (iii) Compute the theoretical isotope pattern from the formula and compare the M+1 (and, where available, M+2) relative intensity with the feature's observed Isotope Distribution; record the agreement and the carbon count implied by the observed M+1 (≈ M+1 % ÷ 1.07), noting that low-abundance features give unreliable isotope patterns. These three axes are computed identically for any adduct. A radical molecular anion [M]•⁻ (or any reading in which the observed m/z is taken as the neutral mass) is treated as admissible **only** when the candidate belongs to a known electron-affinic class (quinones, nitroaromatics, polyhalogenated compounds, strongly conjugated or redox-active species); for all other candidates the radical reading is inadmissible (carried to Stage 5 as a down-weight), because electrospray rarely produces radical anions from ordinary metabolites. This rule both prevents spurious radical/neutral-mass matches and avoids wrongly rejecting genuine radical-anion formers.

**Stage 4 — Deductive co-feature test (the core).** For each surviving hypothesis, predict the m/z of every other ion species the same neutral molecule should produce, by applying every admissible dictionary entry to M: the alternative adducts, the in-source neutral-loss forms, the dimer/multimer forms, and the alternative charge states. For each predicted sibling m/z, search the **complete** quality-controlled feature table (all 2,282 features, annotated or not) for a peak within the mass tolerance **and** within a co-elution window of the query feature's retention time, where the window is set from the chromatographic peak width (e.g. ± one peak width, or a fixed ±0.1 min, whichever is stated). For each co-eluting candidate sibling, read the Spearman rank correlation between its abundance profile and the query feature's from the precomputed per-target correlation vector (look up the matched feature's `m/z`; a predicted sibling absent from the vector is "not assessable", and a predicted sibling whose m/z maps back to the target itself is skipped) and record the coefficient and its p value. A sibling is counted as confirming when it is present, co-eluting, and strongly positively correlated (e.g. ρ ≥ 0.7, or above an empirically chosen threshold reflecting the study's co-elution correlations). Because isotopologues were removed by deconvolution, the isotope axis is taken from Stage 3 rather than from a separate M+1 feature; the co-feature evidence here therefore rests on adduct, in-source-fragment, and charge-state siblings. The number and strength of confirmed siblings is the primary discriminator between hypotheses and is fully adduct-general. One important qualification: when the **target itself fails the reliability filters** (Minimum CV% above threshold or Maximum Abundance below the dataset median), the co-variation axis is underpowered — measurement noise in a low-abundance or high-CV feature attenuates the Spearman correlation (regression dilution), so even a true sibling may fall short of the ρ threshold. In that case an empty co-feature result is **uninformative**, not negative evidence, and must not be weighted against any candidate; the assignment ceiling is then set by the mass, formula, and adduct axes alone.

**Stage 5 — Chemical and chromatographic plausibility weighting.** Down-weight (do not delete) hypotheses whose proposed adduct is inadmissible under the mobile phase (formate, fluoride here) or chemically inconsistent with the compound class (e.g. [M−H]⁻ for a quaternary-amine choline lipid, which ionises as [M+CH₃COO]⁻ or [M−CH₃]⁻). Compare the feature's retention time with the expected BEH Amide HILIC elution window for the candidate's polarity/class (nonpolar lipids early, polar metabolites late); record agreement as supporting and gross disagreement as contradicting. Optionally, if MSᴱ fragments at the feature's retention time positively show class-diagnostic ions (for example a fatty-acyl carboxylate plus a glycerophospho-headgroup ion for a glycerophospholipid, or a hexose neutral loss for a glycosphingolipid), record this as positive support; never use a low MSᴱ Fragmentation Score as evidence against, for the reasons above.

**Stage 6 — Score, rank, and select.** Rank the hypotheses for each peak by a transparent, pre-declared combination of: mass error (smaller is better), isotope-pattern agreement, the count and strength of confirmed co-eluting co-varying siblings (Stage 4, weighted most heavily), adduct admissibility, and retention-time plausibility. Report the ranked list rather than only the top hit; where two compound classes or several isomers remain indistinguishable, report them jointly. The selected annotation is the highest-ranked hypothesis whose evidence meets the level criteria in Stage 7. As an interpretive flag, when the only mass match is a radical/neutral-mass reading, no admissible whole-molecule formula exists at any admissible-adduct neutral mass, and the feature is a low-abundance orphan with no co-eluting siblings, report the feature as a likely in-source fragment or analytical artifact rather than a distinct compound (Schymanski Level 5), so the emptiness is correctly attributed to chemistry rather than to a coverage or threshold limitation.

**Stage 7 — Assign a Schymanski confidence level.** Using the assembled evidence, assign and justify a level (Schymanski et al., 2014):
- Level 1 (confirmed) — retention time and MS/MS match an authentic reference standard analysed under the same method.
- Level 2 — a probable structure from a spectral-library MS/MS match (2a) or from unambiguous diagnostic MS/MS with no competing structure (2b).
- Level 3 — one or more tentative candidates, a compound class, or a substructure, supported by accurate mass, isotope pattern, retention-time plausibility, and/or co-feature evidence, but with the exact structure unresolved (multiple isomers and/or competing formulas).
- Level 4 — an unequivocal molecular formula with no structural assignment.
- Level 5 — exact mass of interest only.
With the present data (no standards, MSᴱ fragmentation that is uninformative at typical abundances, and a deisotoped table) the protocol routinely yields Level 3 or Level 4 annotations; Levels 1–2 are reserved for features subsequently confirmed by targeted MS/MS or standards.

**Stage 8 — Report.** For each peak record: observed m/z, retention time, charge, peak width, Maximum Abundance, Minimum CV%; the ranked candidate hypotheses with formula, adduct, mass error (ppm), isotope agreement, and the list of confirmed co-eluting co-varying sibling ions with their ρ; the adduct-admissibility and retention-time-plausibility judgements; any positive MSᴱ diagnostic ions; and the assigned Schymanski level with a one-line justification. State the analyst-validation status (Accepted? flag) and whether the feature passed the reliability filters.

## Worked example (illustrative output format, not part of the method)

The following is an illustration, not a normative step. The numbers below are specific to m/z 708.53613 and must not be carried over to any other peak; every value (candidates, siblings, isotope figures, Schymanski level) is re-derived per peak by running Stages 1–8. The narrative is followed by the filled Stage 8 report template, which fixes the output format used for every peak.

Applying Stages 1–8 to feature 4.56_708.5361m/z: it is a singly-charged, analytically reproducible but low-abundance feature (Minimum CV% 18.7; Maximum Abundance ≈ 579, ~24th percentile and below the dataset median; peak width 0.27 min). Candidate hypotheses from both the Progenesis output and the broad 10-ppm search reduce, after the mass/charge checks, to a PE plasmalogen (C₄₁H₇₈NO₇P) and a hexosylceramide/cerebroside (C₄₁H₇₇NO₉), each as the in-source [M−H₂O−H]⁻ ion (+3.3 and −8.1 ppm respectively). The deductive co-feature test on the quality-controlled feature table finds no co-eluting intact precursor [M−H]⁻ (predicted 726.547), no co-eluting acetate adduct, and no co-eluting charge-state sibling; the observed isotope pattern (M+1 ≈ 24%) under-represents the ~46% expected for a 41-carbon formula, though the feature's low abundance limits the reliability of that comparison. MSᴱ fragmentation is uninformative at this abundance. The annotation is therefore reported as a tentative compound-class assignment — putative PE plasmalogen via [M−H₂O−H]⁻, cerebroside not excluded — at **Schymanski Level 3**, unconfirmed, with the biological interpretation resting on the statistical association rather than a firm chemical identity. The same procedure, with the same reference tables and tolerances, is applied unchanged to every other peak of interest.

The same result expressed in the fixed Stage 8 report format (this is the output shape to reproduce for every peak):

```
PEAK
  Feature label            : 4.56_708.5361m/z
  Observed m/z             : 708.53613
  Retention time (min)     : 4.56
  Charge                   : 1
  Chromatographic width    : 0.27 min
  Maximum Abundance        : 579  (below dataset median ~2,420 -> reliability flag: low abundance)
  Minimum CV%              : 18.7  (<=30% -> passes reproducibility filter)
  Observed isotope (M+1)   : 24.1%  (low-abundance: isotope axis down-weighted)

RANKED CANDIDATES
  1. PE plasmalogen        formula C41H78NO7P   adduct [M-H2O-H]-   mass error +3.3 ppm
       isotope agreement   : M+1 obs 24.1% vs theo ~46% -> poor, but abundance below median so not weighted
       12 indistinguishable positional/regioisomers (PE P-/O-36:2 sum composition)
  2. Cerebroside B (HexCer) formula C41H77NO9    adduct [M-H2O-H]-   mass error -8.1 ppm
       isotope agreement   : not weighted (low abundance)

CONFIRMED CO-ELUTING, CO-VARYING SIBLINGS (rho>=0.70, within +-0.10 min of RT 4.56)
  none
    - [M-H]- precursor 726.547        : absent from the 2,282-feature list (not detected)
    - [M+CH3COO]- acetate adduct      : absent
    - [M+Cl]- (762.52)                : present but not co-eluting (RT 4.90) -> not counted
    - [M-2H]2- / [2M-H]- siblings     : absent
    => orphan feature: no corroborating adduct, in-source-fragment, or charge-state sibling
       (note: target fails the low-abundance reliability flag, so this empty co-feature
        result is uninformative rather than negative evidence; the Level-3 call rests on
        the admissible adduct and class-plausible RT, not on the absence of siblings)

PLAUSIBILITY
  Adduct admissibility     : [M-H2O-H]- admissible (in-source loss) under ammonium acetate
  Retention-time class     : RT 4.56 min consistent with a glycerophospholipid on BEH Amide HILIC
  MSe fragmentation        : Fragmentation Score ~0.02; uninformative at this abundance (not used to reject)

ASSIGNMENT
  Schymanski level         : 3 (tentative compound class)
  Reported identity        : putative PE plasmalogen via [M-H2O-H]- ; cerebroside (HexCer) not excluded
  Justification            : favoured molecular formula + admissible adduct + class-plausible RT, but
                             multiple isomers and a competing class, no co-varying sibling ion, and no
                             usable MS/MS -> above Level 4 (candidate structures named) and below Level 2
                             (no library/diagnostic MS/MS); not analyst-Accepted
  Analyst-validation flag  : not Accepted
```

## Parameters to declare for reproducibility

Report the mass tolerance (ppm) for the candidate search and the sibling search; the co-elution retention-time window and how it is derived from peak width; the Spearman correlation threshold and significance level for counting a confirming sibling; the QC Minimum CV% and Maximum Abundance thresholds for the reliability filter; that the per-target correlation vector was computed over the 154 study samples; the full adduct dictionary used (with z, n, Δ for each entry) and which adducts were treated as inadmissible for the mobile phase; the compound-class retention-time windows; the databases and versions searched and the element ranges/RDBE limits of the fallback formula generator; and the scoring order used in Stage 6. Default values for all of these are fixed in the "Experimental conditions", "Co-variation engine: precomputed correlation vector", and "Fixed numeric parameters and reference tables" sections above. All predictions are deterministic functions of these declared inputs, so the workflow is exactly reproducible and extends to any additional adduct by adding a single dictionary entry.
